# Metamorphosis of COVID-19 Pandemic

**DOI:** 10.1101/2020.05.17.20073189

**Authors:** Farhan Saif

## Abstract

We show phase-wise growth of COVID 19 pandemic and explain it by comparing real time data with Discrete Generalized Growth model and Discrete Generalized Richard Model. The comparison of COVID 19 is made for China, Italy, Japan and the USA. The mathematical techniques makes it possible to calculate the rate of exponential growth of active cases, estimates the size of the outbreak, and measures the deviation from the exponential growth indicating slowing down effect. The phase-wise pandemic evolution following the real time data of active cases defines the impact-point when the preventive steps, taken to eradicate the pandemic, becomes effective. The study is important to devise the measures to handle emerging threat of similar COVID-19 outbreaks in other countries, especially in the absence of a medicine.

## Introduction

On December 27, 2019, a hospital in Wuhan, capital of Hubei province, reported the first mysterious, suspected pneumonia cases to the center of disease control (CDC) in the capital. On January 8, 2020 a new coronavirus was identified as the cause of pneumonia that spread all over China in next three weeks. Since then, the novel coronavirus, COVID-19, spread across the globe in the next month. In March the World Health Organization declared it to be a pandemic, that in the absence of a vaccine became a challenge for mankind with a tendency to grow exponentially if no measure is taken to prevent it. The incubation period of COVID-19 can last for a period of two weeks or longer. During the period of latent infection, the disease may still be infectious. The coronavirus can spread from person to person through respiratory droplets and close contact.

Employing social distancing, following patient history case by case, and locking down the province of Hubei, and de-facto quarantining the other provinces by restricting the movement of people China controlled the COVID-19 spread, as reported, in thirty five days successfully with eighty three thousand confirmed active cases. On the other hand similar out break in USA, and Italy remained uncontrolled after fifty days. [1].

In this contribution, based on mathematical modelling, phase-wise evolution of COVID-19 pandemic is defined and explained. Moreover, following real time data of active cases [2, 3, 4] a comparison of the COVID-19 outbreak is provided in four different countries, namely USA, Japan, Italy and China. The study reveals information that is helpful to control and eradicate the pandemic in other countries especially in the absence of a vaccine.

In the presence of an almost consistent growth of young population and large old population, the population pyramid in the USA, Italy and Japan have close resemblance [5], as shown in figure 1. In USA, Italy and Japan the population between 15 and 65 years is 64%, 61.4% and 63%, respectively, whereas the above 65 years of age population is, respectively, 22.41%, 27.1% and 14.79%. However the pandemic has effected the three countries in diversely different pattern.

**Figure 1:**
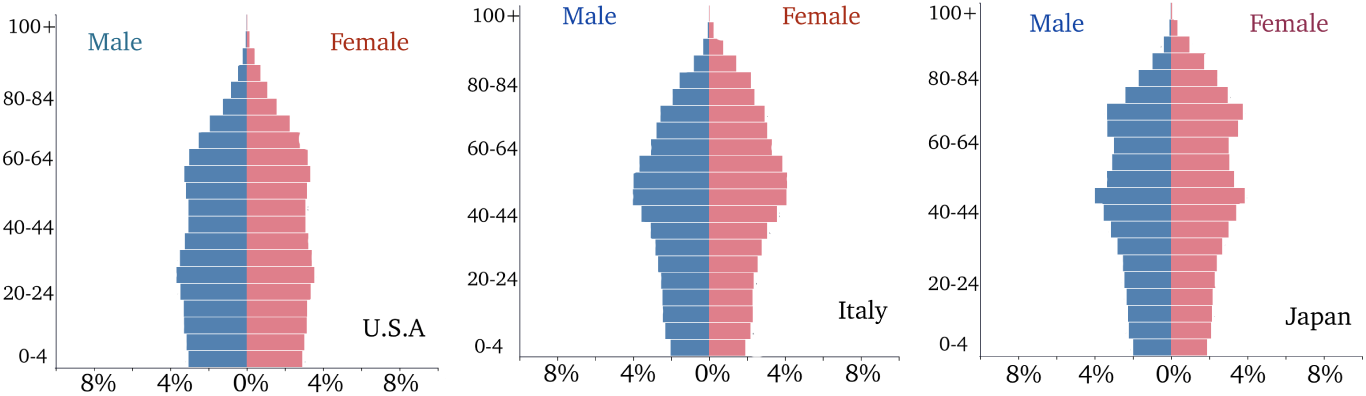
The population pyramid of United States of America (USA), Italy and Japan is provided.

## Following discrete generalized growth model (DG^2^M)

the real-time data of active cases (including infected, recovered and deceased cases) [2, 3, 4], *C_n_*, is compared with the change in active cases, Δ*C_n_*. It is found that logrhythm of change in the active COVID-19 patients per day (log Δ*C_n_*) follows a linear behaviour with the logrhythm of the active patients (log *C_n_*), such that log Δ*C_n_* = *p* log *C_n_* + log *r*. The outbreak follows a certain rate of growth *r* of its spread, whereas another parameter *p* describes the exponent of the spread of the outbreak: it is sub-exponential for 0 < *p* < 1, exponential for *p* =1, and faster than exponential for *p* > 1. We may understand the role of ’*p*’ as, a patient, on average, infects two healthy persons and each of the two new patients effect two healthy persons each and so on — the outbreak expands exponentially. Whereas *r* defines the rate with which these events in exponential growth are taking place in a day. Interestingly a comparison of the real time data of the active patients with numerical results divides the data into two sub-groups:

### First phase COVID-19

First sub-group of the data describes the evolution of COVID-19 pandemic in its early phase, expressed by red-dashed lines in figure 2. For the USA we obtain the red-dashed line for *r* = 0.496 and *p* = 0.925, for Italy, DG^2^M model provides *r* = 0.33 and *p* = 0.95, whereas in case of Japan *r* = 0.427 and *p* = 0.7. The values of *r* in case of USA and Japan are comparable, however smaller in case of Italy, indicating a higher inflow of number of positively tested carriers from the infected countries in USA and Japan as compared to Italy. In contrast a smaller exponent, *p*, value for Japan corresponds to a slower spread of pandemic as compared with the USA and Italy. That is due to a general social distancing awareness in Japan.

**Figure 2:**
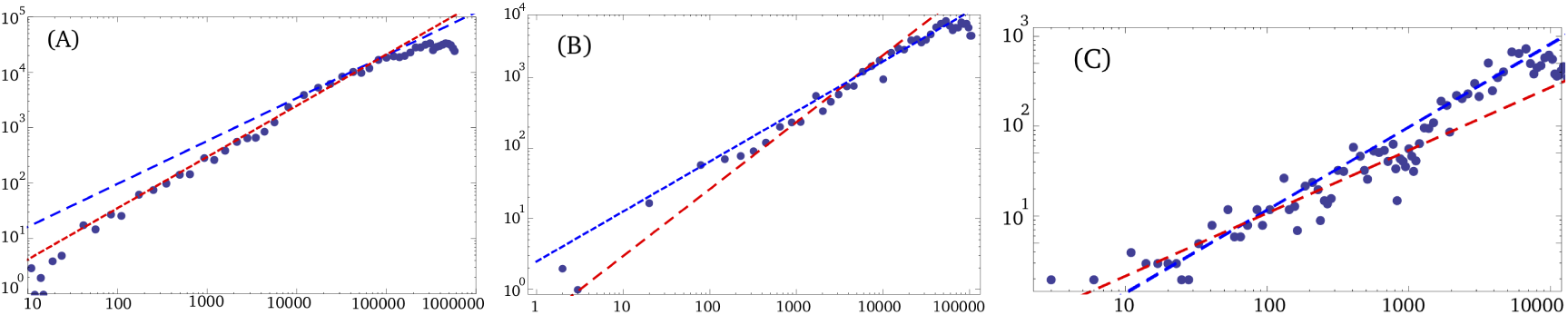
We show log-log plot between the change in the number of active cases per day Δ*C_n_* = *C_n_*_+1_ − *C_n_* and the number of active cases *C_n_* per day for the USA, Italy and Japan: In the panel for the USA we show real time data as dots, following DG^2^M model it makes two straight lines: red dashed-line for *r* = 0.496 and *p* = 0.925, whereas the blue dashed-line for *r* = 2.718, *p* = 0.775. In the panel for Italy we show the real-time data of active cases in Italy making two lines, red dashed-line for *r* = 0.33 and *p* = 0.95, whereas the blue line for *r* = 2.46, *p* = 0.72. In the panel for Japan the red-dashed line plotted by means of DG^2^M has *r* = 0.427, *p* = 0.7, whereas blue line for *r* = 0.169 and *p* = 0.92.

Furthermore, DG^2^M results for Japan show a gradual transition within early phase data, as a straight line is developed by the initial data that has *r* = 1.16, and *p* = 0.55. This gradually transforms to the red-dashed straight line with values *r* = 0.427, and *p* = 0.7. The very early large rate, *r* = 1.16 indicates that a larger number of active cases entered in Japan as foreign travellers from infected countries which had lesser tendency to mingle in the society, keeping a small value for *p* = 0.55.

### Second phase COVID-19

The next sub-group emerges as another linear trend in the real time data following DG^2^M, that appears as the pandemic is slowing down. In case of the USA by employing DG^2^M we get the linear behaviour with *r* = 2.718, *p* = 0.775. For Italy, with the help of DG^2^M, we get *r* = 2.46, *p* = 0.72, and in case of Japan we have *r* = 0.169 and *p* = 0.92. The very small *r* value for Japan and the exponent *p* comparable with USA and Italy indicates a swift approach on day-to-day basis that restricted the number of patients comparatively to lower value so that after eighty days without locking down in the country the number of active cases are very low.

## The Discrete Generalized Richard Model (DGRM)

provides the pandemic evolution after the second phase of COVID-19. The DGRM is valid when the DG^2^M breaks beyond the point of inflection. The DGRM reads as 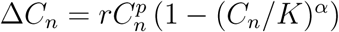, where a shows the deviation from the exponential growth and *K* is the scaling number related to the size of the outbreak or maximum number of active cases. The values of *r* and *p* come from DG^2^M that defined the second phase of COVID-19. Together with the values of *r* and *p* obtained from DG^2^M, the number of active cases in USA show an overlap with DGRM results for *α* = 0.78, and *K* = 1250, 000. We show the overlap of the actual number of active cases and the corresponding DGRM results for *C_n_* versus *n* in figure 3. In case of Italy we have *α* = 1.25 and *K* = 210000 whereas in case of Japan we simulate the real time data by means of *α* = 1.7 and *K* = 17000, together with corresponding *r* and *p* values from DG^2^M.

**Figure 3:**
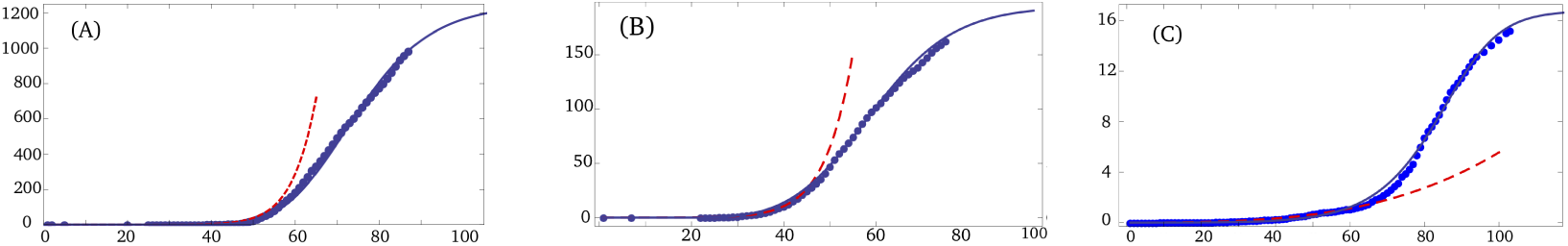
We plot the number (in thousands) of active cases *C_n_* versus the number of days, *n*, in the alphabetical order for USA (A), Italy (B) and Japan (C). The red dashed-line is obtained by DG^2^M, describes the first phase of the pandemic. The blue dashed-line comes from DG^2^M, whereas DGRM result is shown as blue line. Together with the values of *r* and *p* obtained from DG^2^M, (A) in case of the USA we have *α* = 0.78, and *K* = 1250, 000, (B) in case of Italy we have *α* = 1.25 and *K* = 210000. (c) In case of Japan, *α* = 1.7 and *K* = 17000.

An excellent match between numerical plots and real time data confirms the parametric evolution. From the figure 3 we note that the discrete generalized growth model (DG^2^M) show the gradual increase at first phase that matches very good with the real time data of active patients in USA, Japan and Italy. The deviation from unrestricted exponential growth comes from DGRM, that indicates a positive effect of the steps taken by the countries, for example in case of Italy strict lock down, restricting intercity train and bus travel, closure of education sector, and limiting unnecessary travels as the positively tested cases reached around ten thousands. However the preventive measures taken in Japan appear in a very controlled evolution in the DG^2^M model, scaling down the increase on every day basis without locking down the country, and keeping the business sector open, hence effecting the economic activities to the least.

### Developing situation

The phase-wise evolution is seen as making two straight lines as shown in figure 2. The point of intersection of the two lines or the point of impact may be associated with the efficacy of the measures to limit the pandemic in each country as a function of time. In case of China, the intersection corresponds to a general lock down in most of the provinces of China on January 26,2020, as soon as the reported cases were around one thousands.

The real time data from Italy show the point of impact when the *C_n_* value is around 10000. Italy applied serious lock down and restrictions after almost an order of magnitude increase in the number of active cases, that is around 10000. In the night between 7 and 8 March, the Italian government approved a decree to lock down Lombardy and 14 other provinces in Veneto, Emilia-Romagna, Piedmont and Marche, involving more than 16 million people.

In case of USA the point of impact appeared after another order of magnitude increase, that is seen as a slower reaction to combat the pandemic, for instance, due to lack of testing kits. By mid-March, the USA had tested 125 people per million of their population, which was lower than several other countries. On March 27, the U.S. President Trump signed the stimulus bill that approved $2 trillion to fight against COVID-19. The number of active patients had already reached one hundred thousands by that day.

In case of Japan, however, the point of impact appeared as the number of cases were one hundred, as shown in figure 2, that implies a very quick response of the country to the pandemic. For example, on 16 February, Abe convened the government’s first Novel Coronavirus Expert Meeting at the Prime Minister’s Office to draft national guidelines for COVID-19 testing and treatment.

### Expansion of the COVID-19 pandemic

The logrhythmic evolution, in figure 4, expresses the size of the outbreak in USA, Italy, Japan, and China. The numerical calculations are based on the DGRM and show very good agreement with the real time reported data of the pandemic, it is expected that in USA in the presence of strict measures the coronavirus pandemic may effect around 1.25million people before the situation approaches to stability, and 0.21 million in Italy. Whereas for Japan the estimated number of active patients is around sixteen thousands by the time the pandemic is appreciably eradicated, that is possible by mid of May.

**Figure 4:**
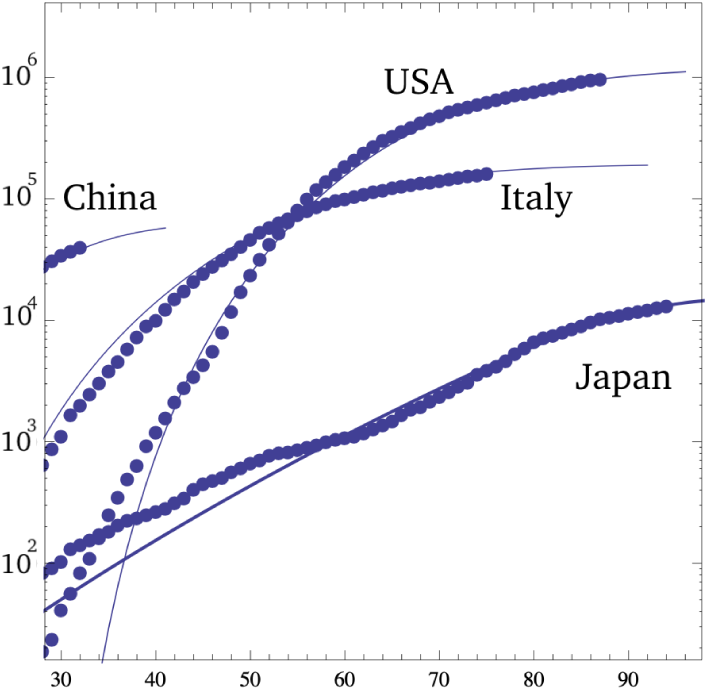
Along the vertical axis we show the logrhythm of number of active cases of COVID-19, *C_n_*, and the number of days *n* are along the horizontal axis. The blue dashed-lines are obtained by DGRM whereas the blue dots represent the real-time data. The values of the parameters are the same as given in figure 2, and figure 3.

## Conclusions

In the analysis the phase-wise growth of COVID 19 pandemic is defined and explained based on DG^2^M and DGRM models. Furthermore the study explains the exponential growth, provides the rate of growth, and estimates the size of the outbreak, and measures the deviation from the exponential growth indicating slowing down effect. The phase-wise pandemic evolution following the real time data of active cases indicates the effectiveness of preventive steps taken in these countries. It is shown that the preparedness of Japan in dealing with the COVID-19 pandemic is the highest, that makes it promising to initiate the 2020 Olympics in the next year. The study is important to devise the measures to handle the emerging threat of similar outbreaks in other countries. Moreover the research work leads us to identify suitable model needed to handle eradication of COVID-19 pandemic in already effected countries in the absence of tested medicines.

## Data Availability

data is taken from Wikipedia

## 1 Acknowledgement

The authors thank for Giulio Casati, Edson D. Leonel, Dieter Suter, Irene Marzoli, Shinichi Watanabe for the useful comments on the manuscript.

## References

[1] F. Saif, Signature of State measures on the COVID-19 Pandemic in China, Italy, and USA, medRxiv. 2020. Preprint at: https://www.medrxiv.org/10.1101/2020.04.08.20057489

[2] https://en.wikipedia.org/wiki/2020_coronavirus_pandemic_in_the_United_States

[3] https://en.wikipedia.org/wiki/2020_coronavirus_pandemic_in_Italy/

[4] https://en.wikipedia.org/wiki/2020_coronavirus_pandemic_in_Japan

[5] https://www.populationpyramid.net

